# Perioperative Hyperoxia and Delirium after On-pump Cardiac Surgery: A Mediation Analysis

**DOI:** 10.1101/2022.06.07.22276112

**Authors:** Kwame Wiredu, Stefana Voicu, Heba Naseem, Ariel L Muller, Myles D Boone, Scott A. Gerber, Shahzad Shaefi

**Affiliations:** Geisel School of Medicine at Dartmouth, Hanover NH; Department of Anesthesia, Critical Care and Pain Medicine, Beth Israel Deaconess Medical Center, Boston MA; Department of Anaesthesia, Critical Care and Pain Medicine, Harvard Medical School, Boston MA; Department of Anesthesiology, Dartmouth-Hitchcock Medical Center, Lebanon, NH; Dartmouth Cancer Center, Geisel School of Medicine at Dartmouth, Lebanon, NH

**Keywords:** biomarkers, cardiopulmonary bypass, delirium, neuroinflammation, plasma

## Abstract

**Background:** Neurologic and neurobehavioural complications are common after cardiac surgery with cardiopulmonary bypass (CPB). Exposure to the artificial bypass surface, conversion to laminar flow and hypothermia likely contribute to systemic inflammation observed after CPB. To ensure adequate systemic oxygenation, the CPB patient is often exposed to supraphysiologic levels of oxygen. Relative to normoxia, perioperative hyperoxia during CPB has not been shown to impact neurocognition in the long-term. Whether this holds true for the immediate post-operative neurocognitive function is the question of this nested case-control study.

**Methods:** 46 age-and sex-matched subjects, aged ≥65 years, selected for this study were randomized to receive normoxia or hyperoxia during CABG with CPB in the parent trial. Levels of four neuroinflammatory biomarkers (S100B, ENO2, CHI3L1, UCHL1) were measured at baseline and at post-bypass. Baseline neurocognition was established with the Montreal Cognitive Assessment tool and patients were assessed on each post-operative day for delirium using the confusion assessment method. Mediation analyses was conducted for the conditional effect of perioperative oxygen treatment on the occurrence of delirium, assuming mediation effect from change in biomarker levels.

**Results:** 26 subjects (*n* = 12) demonstrated delirium. Of the four biomarkers, only S100B levels were differentially abundant post-bypass regardless of treatment (8.18 versus 10.15pg/mL, *p* value < 0.001). We found significant direct effects of treatment on the occurrence of delirium (effect size = -2.477, *p* = 0.004). There was no statistically significant mediating effect by S100B.

**Conclusion:** While perioperative hyperoxia may not be associated with neurocognitive dysfunction in the long-term, its immediate effects may contribute significantly to the occurrence of post-operative delirium. Taken together, our findings suggest a dose-response-time relationship between hyperoxia and neurocognitive function.

## Introduction

The effects of perioperative hyperoxia on myocardial damage, acute kidney injury and long-term neurocognitive dysfunction are well documented,[1-6] but the impact on the immediate post-operative neurocognitive function is less well-characterized.[7] Globally, over 1.25 million patients undergo cardiac surgery on cardiopulmonary bypass (CPB) each year.[8] Perioperatively, the CPB population are often exposed to supraphysiologic concentrations of oxygen.[9] This practice is premised on the primary goal of maintaining end-organ perfusion as hypothermia, microcirculatory heterogeneity and interstitial fluid shifts during CPB all contribute to poor tissue oxygenation.[10]

At the tissue level, hyperoxia is beneficial in the ischemic preconditioning of the myocardium, reduces overall gas microemboli and provides significant oxygen reserves in the event of interrupted ventilation.[11-13] In fact, the ability to monitor regional cerebral oxygenation in real-time has provided unequivocal evidence linking cerebral desaturations during CPB to worse clinical outcomes,[14-17] further emphasizing the need for higher oxygen concentrations. On the other hand, hyperoxia may also trigger vasoconstriction that further compromises perfusion, may instigate inflammation and worsen ischemia-reperfusion injury through increased oxidative stress.[18-21] Notwithstanding these, the prevailing observation is that hyperoxia during CPB is not associated with poor long-term neurocognitive outcomes.[2, 5, 22]

Delirium is etiologically heterogeneous and lower pre-surgical cognitive reserve is a recognized risk factor.[23-25] The extent to which perioperative oxygen treatment modifies the occurrence of delirium in a typical CPB demographic with suboptimal baseline neurocognition, however, remains largely unknown. In this nested case-control study, we hypothesized that peri-operative oxygen administration significantly impacts the occurrence of post-operative delirium. We estimated the conditional effect of perioperative hyperoxia in an elderly cohort who underwent cardiac surgery on CPB.

Further, we measured a panel of neuroinflammatory markers and ascertained their possible role in mediating the hyperoxia-delirium relationship. Finally, we proposed a conceptual model regarding the interaction between baseline neurocognition and perioperative hyperoxia as they relate to post-operative delirium.

## Methods

### Study design and Ethics approval

Subjects in this nested case-control study were selected from the parent clinical trial[5, 26] that examined the effects of intra-operative oxygen therapy on neurocognitive outcomes among cardiac surgical patients at the Beth Israel Deaconess Medical Center, Boston MA (Trial registration number NCT02591589, principal investigator: Shahzad Shaefi, registration date: October 29, 2015). Subjects were enrolled between July 2015 and July 2017, and all patients provided informed consent. Institutional review board (IRB) approval 2014P000398/33 was amended for the purposes of this current study on 09/17/2021 by the Committee on Clinical Investigations (CCI) at the BIDMC. **Figure 1** summarizes the design of the current study.

**Figure 1:**
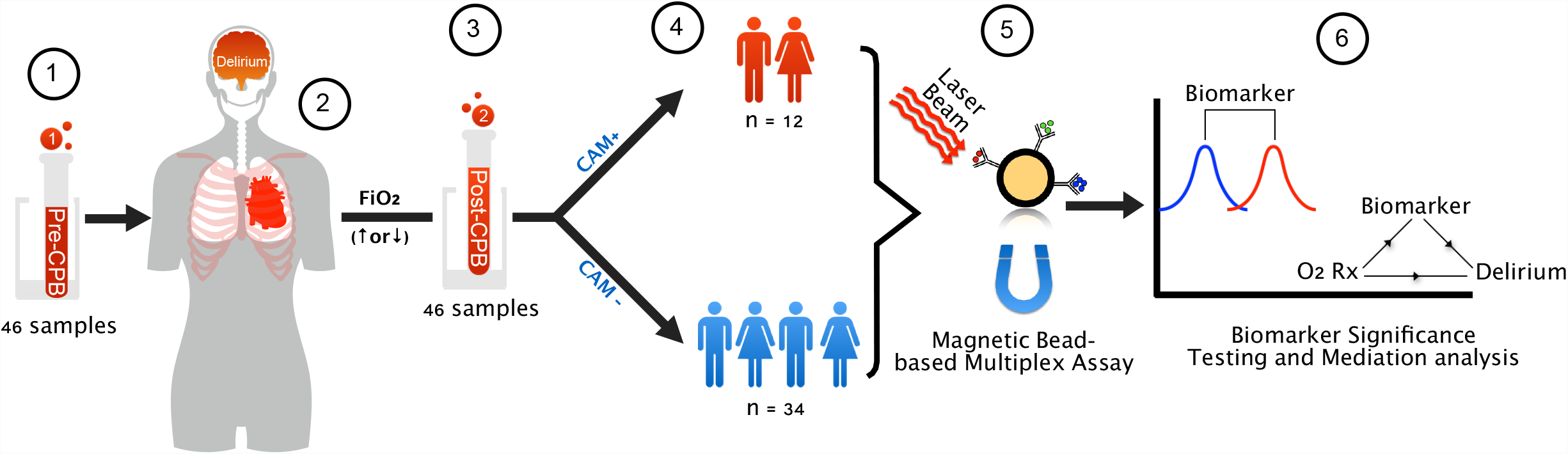
Study design and schematic illustration of the nested case-control study. Baseline plasma samples (**1**) were collected prior to CABG on CPB during which patients were randomized to normoxia or hyperoxia treatment groups (**2**). Plasma samples were collected at post-bypass (**3**). Following surgery, patients were followed with daily assessments for delirium using CAM (**4**). Biomarker concentrations in plasma were measured in duplicates by bead-based multiplex assay (**5**), for subsequent data analyses (**6**). CAM: confusion assessment method; CPB: cardiopulmonary bypass; FiO_2_: fraction of inspired oxygen; O_2_ Rx: intra-operative oxygen treatment

### Patient population, Exclusion and Inclusion Criteria

Details of enrollment, exclusion criteria and treatment allocation are published previously.[5, 26] Briefly, eligible participants included patients 65 years or older who were booked for elective CABG requiring CPB. Neurocognitive assessment was achieved using the telephonic Montreal Cognitive Assessment (tMoCA) as the primary endpoint. Post-operatively, subjects were assessed daily for delirium as a secondary endpoint using the confusion assessment method (CAM). Patients were excluded if they were undergoing emergent CABG, if they required single-lung ventilation, CABG without CPB, intraoperative balloon counter-pulsation or mechanical circulatory support. Subjects with MoCA scores below 10 were also excluded.

### Sample size calculation

Because quantitative studies on the selected biomarkers in the context of delirium are largely unexplored, Cohen’s estimation of effect size[27] was used to determine the optimal sample size. Here, we used a standardized effect size by Cohen’s estimation, *d* = 0.8 for a paired design that provides a statistical power of 80% at a significance level of 0.05. Further, delirium cases were also matched to non-delirium controls at approximately 1:3 ratio, to a total of 46 subjects in the current study.

### Conduct of Study and Biomarker Measurements

Whole blood samples at baseline and post-bypass (P-BP) were collected into 4mL EDTA-treated tubes (BD Diagnostics) and centrifuged immediately. Resulting plasma was stored at -80ºC until analyses. Limits of detection, limits of quantification and linearity of biomarker signal were established using serial dilutions of patient samples and laboratory standards. Biomarker measurements were made using a custom R&D Human Premixed Multi-Analyte Panel, a magnetic bead-based multiplex assay (Catalog Number: LXSAHM-04, Lot Number: L140030). Analyte concentrations were determined by a 5-parameter logistic (PL) regression computed from the standard curves. All biomarkers were measured in duplicates. For quality control, intra-assay variability was assessed at a cut-off of 20%.

### Statistical analyses

Descriptive statistics are presented as mean (standard deviations) or count (proportion) for continuous and categorical variables, respectively. Biomarker concentrations, which were assayed in duplicates, were summarized as averages and compared between groups using Student’s *t*-test (paired, unequal variance). Structural equation modeling (SEM) was employed to ascertain the conceptual and statistical mediation models, hypothesizing a direct effect of oxygen treatment on delirium outcome, and an indirect (mediation) effect via the measured biomarkers. (**Figure 4A** and **Supplemental Figure 1**). Mediation analysis was then performed to estimate the total, indirect and direct effects of intraoperative oxygen treatment on the occurrence of delirium. Here, the direct effect of perioperative oxygen treatment was computed as the change in the odds of developing delirium in patients receiving hyperoxia versus the odds of developing delirium in patients receiving normoxia, when baseline neurocognition is fixed (i.e., holding tMoCA scores constant). Average tMoCA score was also defined as the arithmetic mean of tMoCA scores for subjects in each arm of this nested cohort. Significance of the mediation effects were computed by bootstrapping.[28] All analyses were performed in R environment for statistical computing, v4.1.1.[29] at a significance level ⍰ = 0.05.

## Results

26% of subjects (n = 12) in the nested cohort demonstrated delirium postoperatively. At baseline, delirium cases and non-delirium controls were matched by age, sex and race (**Table 1**). Details of subjects’ comorbidities and preoperative medications are reported previously.[5] Intraoperatively, there was no statistically significant differences in aortic cross-clamp or cardiopulmonary bypass times between cases and non-cases. Baseline neurocognition, as measured by the Montreal Cognitive Assessment (MoCA) tool, was generally low in this cohort, and significantly lower among cases relative to controls (*p* = 0.02). Of the non-delirium controls, 21% (*n* = 7) met the criteria for subsyndromal delirium.

**Table 1:** Baseline characteristics of study participants

|  | Case<br>(N = 12) | Non-case<br>(N = 34) | p-value |
| --- | --- | --- | --- |
| **Demographics** |  |  |  |
| Age (years)<br>*mean (SD)* | 74 ($\pm 6.4$) | 70 ($\pm 4.3$) | 0.07 |
| Male sex<br>*count (%)* | 9 (75%) | 27 (79%) | 1 |
| BMI (*count (%)*) |  |  | 0.12 |
| Underweight | 1 (8%) | 1 (3%) |  |
| Normal | 2 (17%) | 7 (21%) |  |
| Overweight | 1 (8%) | 13 (38%) |  |
| Obese | 8 (67%) | 13 (38%) |  |
| White race<br>*count (%)* | 10 (83%) | 34 (100%) | 0.11 |
| **Baseline neurocognitive assessment** |  |  |  |
| Pre-operative MOCA<br>*mean (SD)* | 15 ($\pm 2.7$) | 17 ($\pm 2.1$) | 0.02 |
| **Intraoperative** |  |  |  |
| Hyperoxia<br>*count (%)* | 4 (33%) | 19 (56%) | 0.31 |
| Cardiopulmonary bypass time (mins)<br>*mean (SD)* | 79 ($\pm 17$) | 81 ($\pm 24$) | 0.83 |
| Aortic Cross-clamp time (mins)<br>*mean (SD)* | 66 ($\pm 14$) | 67 ($\pm 21$) | 0.85 |

| Postoperative |  |  |  |
| --- | --- | --- | --- |
| Delirium severity score<br>*mean (SD)* | 8.9 ($\pm 3.1$) | 3.8 ($\pm 2.0$) | < 0.01 |
| Sub-syndromal delirium<br>*count (%)* | 0 (0%) | 7 (21%) | 0.22 |

†4 subjects have missing data on CPBT and XCT, and 1 subject missing data on delirium severity

We measured four biomarkers well-documented to be markers of neuro-inflammation. For quality control, we set an intra-assay variability cut-off of 20%. Biomarker measurements with 20% coefficient of variation (%CV) or higher between duplicate runs were removed from all downstream analyses, although including them did not change results of our analyses. Levels of ubiquitin carboxyl-terminal hydrolase isozyme L1 (UCHL1) in all samples were below the limits of quantification (**Supplemental table 1**).

Of the remaining biomarkers, only S100B levels were significantly higher at post-bypass relative to baseline levels (*p* value < 0.001, **Figure 2**). We found no significant differences in S100B levels by sex or body mass index (BMI) at baseline or post-bypass (**Table 2**). Stratified analyses also showed that the absolute increase in S100B levels is not confounded by sex, BMI, patient outcomes or intra-operative oxygen treatment (**Table 2, Supplemental figure 2**). We, however, observe that for patients with the longest CPB times (> 141 mins), the absolute change in S100B levels was not significant (*p* = 0.076, **Table 2**).

**Table 2:** Stratified analysis of baseline to post-operative change in S100B levels

|  | Baseline S100B (pg/mL) | Postop S100B (pg/mL) | *t*-statistic | *p*-value |
| --- | --- | --- | --- | --- |
| Overall |  |  |  |  |
|  | 8.18 | 10.15 | 7.68 | <0.001* |
| Stratified by outcome |  |  |  |  |
| Case | 8.66 | 10.70 | 4.291 | 0.004* |
| Non-case | 8.05 | 9.99 | 6.397 | <0.001* |
| Stratified by treatment |  |  |  |  |
| Hyperoxia | 8.08 | 10.10 | 5.015 | <0.001* |
| Normoxia | 8.30 | 10.21 | 6.057 | <0.001* |
| Stratified by BMI<sup>§</sup> |  |  |  |  |
| Normal | 8.17 | 10.61 | 4.125 | 0.004* |
| Overweight | 7.81 | 10.03 | 5.907 | <0.001* |
| Obese | 8.47 | 10.01 | 3.824 | 0.002* |
| Stratified by Sex |  |  |  |  |
| Female | 9.09 | 10.65 | 3.372 | 0.015* |
| Male | 7.96 | 10.03 | 6.922 | <0.001* |
| Stratified by CPB Time |  |  |  |  |
| < 52 mins | 8.47 | 10.35 | 3.675 | 0.006* |
| 52 – 112 mins | 8.23 | 9.95 | 3.267 | 0.014* |
| 112 – 141 mins | 7.29 | 9.72 | 6.227 | <0.001* |
| > 141 mins | 8.94 | 10.24 | 2.083 | 0.076 |

\*: statistically significant difference at $\alpha \leq 0.05$
§: Underweight category ( $n = 2$ ) is underpowered for parametric testing

**Figure 2:**
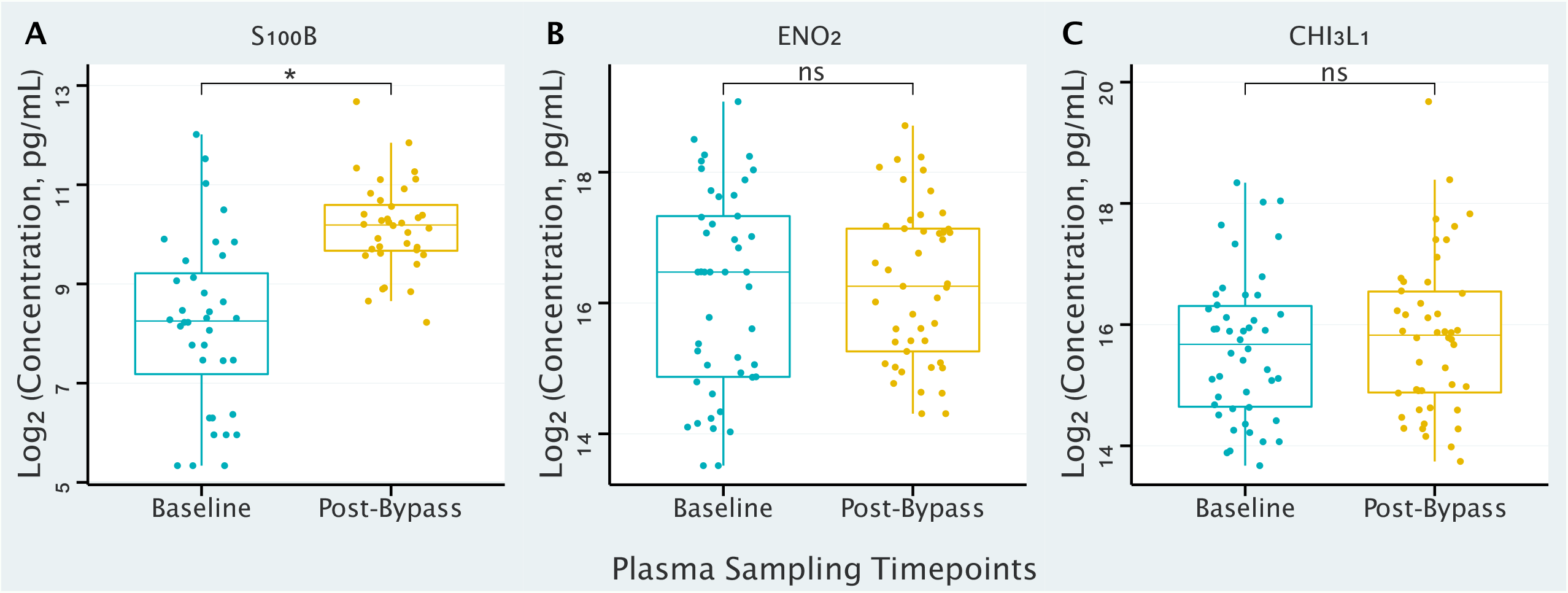
Comparison of neuro-inflammatory biomarker levels. Boxplots show the quantile distribution (median and inter-quantile ranges) of each biomarker, and a comparison of biomarker levels at baseline and at post-bypass. Statistical tool used is the Student’s *t*-test. Asterisks (*) represent statistically significant difference (i.e., *p* value < 0.05); *ns*: not significant; S100B: protein S100-B; ENO_2_: gamma-enolase; CHI3L1: chitinase-3-like protein 1.

There were significant conditional and total effects of intra-operative oxygen treatment on the occurrence of delirium (effect estimate = -2.477, *p* = 0.004, **Tables 3** and **4**). We, however, did not find post-bypass changes in S100B levels to mediate the oxygen-delirium relationship (indirect effect = 0.002, *p* = 0.584, **Table 3** and **Figure 3**).

**Table 3:** Conditional effects and regression coefficients of predictors in mediation modeling

| Variables | Predictors | Label | $\beta$ | CE | SE | $p$ |
| --- | --- | --- | --- | --- | --- | --- |
| $\Delta(S100B)$ | Treatment | $a_1$ | 0.412 | 1.507 | 4.638 | 0.745 |
| $\Delta(S100B)$ | tMoCA | $a_2$ | -0.134 | -0.113 | 0.155 | 0.464 |
| $\Delta(S100B)$ | tMoCA * Treatment | $a_3$ | -0.364 | -0.074 | 0.260 | 0.776 |
| *Delirium* | Treatment | $c_1$ | -0.724 | -2.477 | 0.862 | 0.004* |
| *Delirium* | tMoCA | $c_2$ | -0.138 | - 0.110 | 0.042 | 0.009* |
| *Delirium* | tMoCA * Treatment | $c_3$ | 0.682 | 0.130 | 0.050 | 0.009* |
| *Delirium* | $\Delta(S100B)$ | B | 0.002 | 0.001 | 0.041 | 0.971 |

**Table 4:** Causal Mediation Analysis

| | Estimate | 95% CI | $p$ value |
| --- | --- | --- | --- |
| ACME | 0.00 | 0.00 – 0.02 | 0.584 |
| ADE | -0.08 | -0.11 – -0.02 | 0.013* |
| Total effect | -0.07 | -0.12 – -0.02 | 0.016* |
| Proportion Mediated | -0.03 | -0.37 – 0.08 | 0.596 |

**Figure 3:**
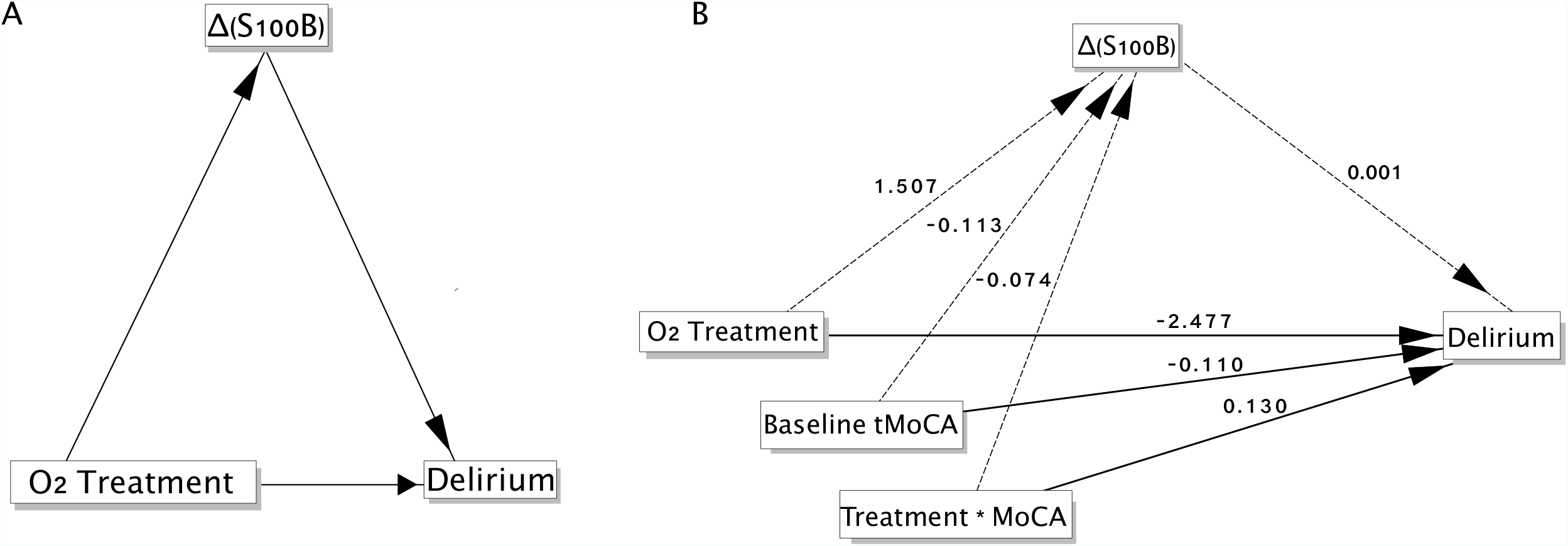
Mediation modeling and analyses. **A**: Conceptual model, and **B**: Statistical model of the relationship between delirium occurrence and intra-operative oxygen treatment, assuming a mediation role by post-bypass increases in S100B biomarker levels. Dashed lines = statistically non-significant; MoCA: Montreal Cognitive Assessment tool used to establish baseline neurocognition; O2: oxygen

## Discussion

We have presented an exploratory, nested case-control study that tested the hypothesis that the amount of intraoperative oxygen administered likely contributes to the occurrence of delirium. We found a significant direct effect between hyperoxia and the odds of developing delirium. We, however, did not find a significant mediating role of neuroinflammatory biomarkers measured in our study. The effects of perioperative oxygen treatment on the occurrence of delirium, however small the effect size may be, reechoes the fact that delirium is etiologically heterogenous with likely many other possible pathomechanistic pathways besides our observation. Despite the paucity of literature on the relationship between perioperative hyperoxia and post-operative delirium, result of our study is consistent with recent studies investigating the matter.[7, 30]

In one of the aforementioned studies, Lopez, Pandharipande [7] monitored the duration of cerebral hyperoxia with oximetry monitors intraoperatively and found that despite the considerable fluctuations in cerebral oxygenation, hyperoxia after a period of hypoxia was most strongly associated with the occurrence of delirium. Authors also observed that in delirium subjects, there was an increase in plasma concentrations of markers of oxidative stress (F_2_-isoprostanes and isofurans), suggesting a possible mediation role. In the remaining study, Kupiec, Adamik [30] established a maximum PaO_2_ cut-off of 33.2 kPa, beyond which post-operative delirium was more likely to occur. Notable differences between these studies and ours is the relatively smaller sample size in our study, the choice of the baseline neurocognitive assessment tool and the frequency of assessments for post-operative delirium.

Traditionally, ascertaining the mediation effect of a variable ***M*** (e.g., biomarker levels) on the relationship between a predictor ***X*** (e.g., peri-operative oxygen treatment) and outcome ***Y*** (delirium) has required that a relationship already exists between ***X*** and ***Y***. This approach is heavily debated, and proposed alternatives suggest that a prior relationship between ***X*** and ***Y***, or the lack thereof, neither proves nor rules out causal associations[31-33]. This was the observation in our study, in which we found that the relationship between ***X*** and ***Y*** is moderated by the interaction of another variable W (baseline neurocognition).

The clinical significance of the post-bypass increases in S100B levels without any significant associations with delirium remains to be determined. This is also the observation by Jönsson, Johnsson [34] and Nguyen, Huyghens [35]. Aptly described by authors as the “controversial significance of early S100B levels after cardiac surgery”, Jönsson, Johnsson [34] measured S100B levels at defined intervals from end of bypass until 48 hours post-surgery, and concluded that the predictive significance of the S100B biomarker is limited.[34] These findings are in sharp contrast to many other studies in which S100B levels were consistently increased in delirium cases regardless of the sampling time after surgery[36-38]. These conflicting findings about the S100B-delirium relationship highlights three possibilities: (1) that plasma levels of S100B do not necessarily reflect CSF levels, (2) the relatively short half-life of S100B (60 to 120 mins) requires that blood sampling is appropriately timed, or (3) that increases in S100B levels only signal neuronal response to the surgical insult, and not because of the occurrence of delirium. With regards to the gap in plasma-to-CSF levels, S100B is also secreted by extra-neuronal tissues (e.g., adipocytes)[39, 40], although the predominant source remains in mature, perivascular astrocytes[41, 42]. To ascertain that the post-bypass increases in S100B levels in our cohort were not confounded by body fat, we performed stratified analyses and found no differences in S100B levels by sex or by BMI.

Further, although there was no association between CPB duration and incidence of delirium, we observe a downtrend in the absolute change in S100B levels with increasing CPB times and found no significant change in S100B levels for subjects with the longest CPB times (> 141 mins). We intimate that this is likely due to the short half-life of the S100B protein, although this hypothesis will require a formal interrogation in future studies.

Our study is, however, without limitations, notable among them is sample size and selection bias. Our relatively small sample size (*n* = 46) did not allow for the statistical adjustment of covariates such as age, sex, BMI in the structural modeling and mediation analyses. We, however, controlled for possible confounding during the study designing stage by matching cases to controls by age, sex, BMI and race (**Table 1**), and do not expect the lack of statistical adjustments to have any significant impact on the strength of associations in our findings. To prevent selection bias, subjects in the present study were selected to reflect the distribution of treatment (hyperoxia versus normoxia) and outcome (delirium case versus non-delirium control) in the parent trial. We acknowledge that our cohort had considerable deficits in neurocognition at baseline. While the ideal choice of controls would be subjects without any baseline deficits, controls are more appropriately sampled from, and in terms of risk, should be representative of the very population that gave rise to the cases being investigated[43, 44]. Nonetheless, we excluded subjects with extremely low tMoCA scores (< 10). In addition, patients’ baseline tMoCA scores were included, and statistically adjusted for, in our models.

There are several mechanisms proposed to underlie the possible neurological damage after cardiac surgery on CPB [45]. In our study, we focused on the neuroinflammatory mechanism as a possible mediator of the exposure-to-outcome relationship. It is likely that our choice of biomarkers, albeit their recognized associations with neuroinflammation, may not be directly involved in the pathogenesis of post-bypass delirium. Given the half-lives and turnover of many inflammatory proteins, it is equally likely that our timing of blood sampling did not permit detection of biomarker level that accurately reflects any possible neuroinflammatory process that may have been at play.

To date, our study remains the only one that has investigated the effects of perioperative hyperoxia on both the immediate and long-term neurocognitive functions [5] in the same cohort of patients. Taken together, the findings that hyperoxia increases the risk of post-operative delirium, yet with no association with long-term cognitive decline, may best be explained by a dose-response-time relationship, although further studies are required to definitely interrogate these hypotheses.

## Data Availability

All data produced in the present study are available upon reasonable request to the authors

## Acknowledgements

Funding for this work was supplied partly by the Burroughs Wellcome training grant to KW and via K08 GM134220 and R03 AG060179 to SS.

## Figure Captions and Table Legends

**Supplemental figure 1:**
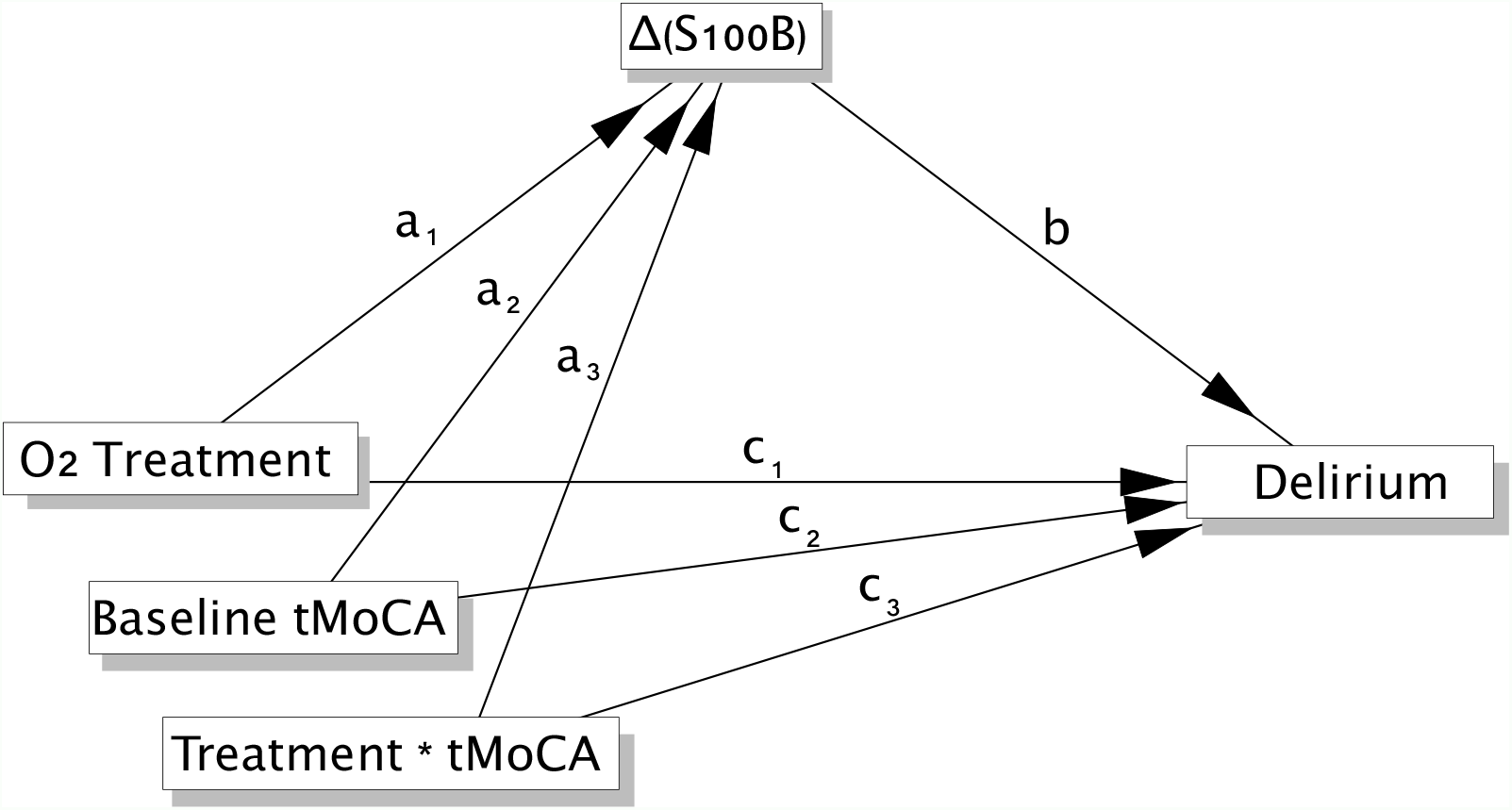
Structural equation model diagram. Delirium is the dependent latent variable; intra-operative oxygen treatment is the independent variable and post-bypass change in S100B biomarker levels is the assumed mediator of the oxygen-delirium relationship

**Supplemental Figure 2.**
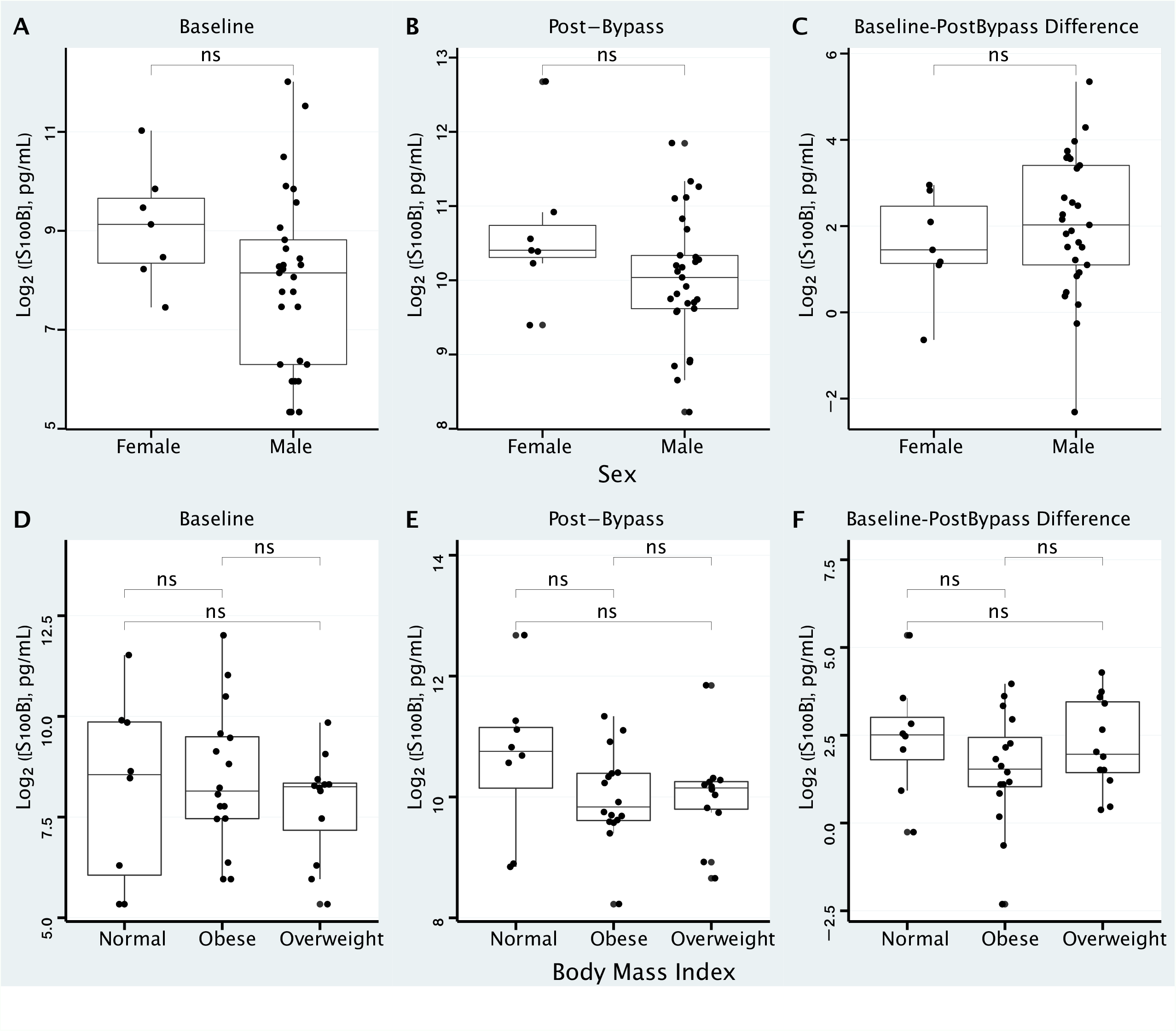

**Supplemental Table 1:** Measured Biomarkers and their Dynamic Range

| Analyte | Dynamic Range (pg/mL) |
| --- | --- |
| Chitinase 3 -like 1 | 438.44 – 106,540 |
| S100B | 40.41 – 9,820 |
| Enolase 2 | 374.44 – 90,990 |
| UCH-L1 | 925.10 – 224,800 |

Table 1: Baseline characteristics of study participants

Table 2: Stratified analysis of baseline to post-operative change in S100B levels

Table 3: Conditional effects and simultaneous regression coefficients of predictors in mediation modeling. β: regression coefficients; CE: estimate of conditional effects; SE: bootstrap standard error; *p*: p values; *: significant association

Table 4: Causal Mediation Analysis. ACME: average causal mediation effects; ADE: average direct effects

## Notes

### Competing Interest Statement

The authors have declared no competing interest.

### Clinical Trial

NCT02591589

### Clinical Protocols

https://clinicaltrials.gov/ct2/show/NCT02591589

### Funding Statement

Funding for this work was supplied by the Burroughs Wellcome training grant to KW, the National Institutes of Health (R01 GM122846) to SAG, and K08 GM134220 and R03 AG060179 to SS.

### Author Declarations

Institutional review board (IRB) approval 2014P000398/33 was amended for the purposes of this current study on 09/17/2021 by the Committee on Clinical Investigations (CCI) at the Beth Israel Deaconess Medical Center, Boston MA

